# Clinical patterns in a neuroimaging-based predictive model of self-reported dissociation

**DOI:** 10.1101/2025.02.27.25323014

**Authors:** Juliann B. Purcell, Boyu Ren, Cori A. Palermo, Zoe A. Bair, Mollie C. Marr, Rebecca L. Modell, Xi Pan, Matthew A. Robinson, Meghan E. Shanahan, Michaela B. Swee, Milissa L. Kaufman, Lauren A. M. Lebois

## Abstract

**Objective:** Assessment of trauma-related dissociation has been historically challenging given its subjective nature and the lack of provider education around this topic. Recent work identified a promising neural biomarker of trauma-related dissociation, representing a significant step toward improved assessment and identification of dissociation. However, it is necessary to better understand clinical factors that may be associated with this biomarker.

**Method:** Participants were 65 women with histories of childhood maltreatment, posttraumatic stress disorder (PTSD), and varying levels of dissociation (e.g., co-occurring dissociative identity disorder, DID). Data were drawn from a previously published work that identified a model predicting Multidimensional Inventory of Dissociation severe pathological dissociation scores on the basis of neural functional connectivity. Here, we conducted a k-means cluster analysis to explore patterns in results of the prediction model. We then investigated differences among the clusters in a range of clinically-relevant variables.

**Results:** The clustering analysis identified four distinct groups. The functional connectivity model best predicted those at the low (cluster 1, 82% PTSD) and high (cluster 3, 86% DID) ends of the self-reported dissociation scale. Cluster 2 also largely included participants with DID (67%), but the predictive model was less accurate for these individuals. Follow up analyses revealed that DID participants in cluster 2 reported lower levels of self-state intrusions, a type of DID-specific dissociation, compared to those in cluster 3.

**Conclusions:** The predictive performance of the functional connectivity biomarker is linked to DID-specific dissociation. This suggests that in the future functional connectivity signatures may improve accurate assessment of DID.

**Clinical Impact Statement:** The present study aimed to examine patterns in a previously identified brain signature of dissociation. We identified two distinct groups of individuals with dissociative identity disorder (DID) who differed in a DID-specific type of dissociation. These findings suggest that the brain signature of dissociation may be linked with DID-specific dissociation and underscore the importance of comprehensively evaluating dissociative symptoms. Dissociative symptoms are difficult to assess because the experience is highly subjective and because many providers do not receive training in dissociation. Objective, brain-based metrics to supplement self-reports would be invaluable in enhancing the assessment and treatment of trauma-related dissociation.

## Introduction

Pathological dissociation encompasses a range of experiences related to the disconnection from one’s internal or external environment and result in significant distress or impairment (American Psychiatric Association, 2022). Although dissociative experiences are transdiagnostic, they are a particularly prominent feature of trauma-spectrum disorders such as posttraumatic stress disorder (PTSD) or dissociative identity disorder (DID) (Dalenberg et al., 2012; Vissia et al., 2016). The clinical relevance of severe pathological dissociation cannot be understated, as prior work strongly suggests it negatively impacts treatment course (Lanius et al., 2012; Price et al., 2014). However, accurately assessing dissociation can be challenging due to the subjective nature of the phenomenon (Lebois, Ross, et al., 2022; Robinson et al., 2024). For example, patients may have difficulty conveying their experiences or may not think to share what are habitual occurrences. Additionally, providers may have difficulty accurately assessing for dissociation given heterogeneity in dissociative experiences among individuals. These conditions, further compounded by a lack of professional education regarding dissociation, lead to poor outcomes and prolonged suffering for patients (Brand et al., 2022; Nester et al., 2024). Objective measures, such as a neural signature of severe pathological dissociation, could be a helpful complement to individuals’ reports, thereby improving recognition, assessment, and, ultimately, treatment outcomes.

Objective measures of severe pathological dissociation will need to account for patterns specific to certain trauma-related disorders, such as PTSD, the dissociative subtype of PTSD, and DID. For example, in PTSD, numbing is often observed, in which the person feels disconnected from and unable to experience their emotions (American Psychiatric Association, 2022). To illustrate, one could think of a military veteran who has difficulty experiencing positive or negative emotions. The dissociative subtype of PTSD involves the presence of depersonalization or derealization, in addition to the standard criteria for PTSD (American Psychiatric Association, 2022). Depersonalization refers to a sense of disconnection from one’s own internal experience or physical body, while derealization refers to a sense of disconnection from the external environment (American Psychiatric Association, 2022). For example, one may report feeling as if they are in a dream anytime they drive after experiencing a traffic collision. Finally, individuals with DID report a wide range of dissociative symptoms. In addition to those already described (i.e., numbing, depersonalization, derealization), individuals with DID also experience a disruption in their sense of a cohesive identity. For example, the intrusion of a thought or emotion into conscious awareness accompanied by a sense that the thought or emotion does not belong to them. Of note, this “not me” experience is not akin to psychosis, as individuals with DID have intact reality testing and are able to recognize that the thought *must* belong to them, despite the sense that it does not (Brand et al., 2009; Brand & Loewenstein, 2010; Dell, 2006; Renard et al., 2017). Although these manifestations of dissociative experiences may seem wholly separate, they are theorized to serve a common function. Dissociation provides a sense of psychological distance from an experience, thereby helping the individual cope with overwhelming pain, trauma, or stressors (Brand et al., 2006; Brenner, 2001; Liotti, 1992). Thus, dissociation is an effective coping mechanism in certain situations, but can become disruptive in other circumstances (Lebois, Ross, et al., 2022; Loewenstein, 2020; Purcell et al., 2024).

In light of the distress and impairment dissociative symptoms can cause and the challenging nature of assessment (Lebois, Ross, et al., 2022; Robinson et al., 2024), the need for objective measures of dissociation is clear. Recent work has begun to explore the identification of objective neural signatures, or biomarkers, of dissociation across a range of modalities (e.g., functional magnetic resonance imaging [fMRI], structural magnetic resonance imaging [MRI], positron emission tomography) (Frewen & Lanius, 2006; Lebois, Kumar, et al., 2022; Nicholson et al., 2019; Reinders et al., 2016; Roydeva & Reinders, 2021). Specifically, studies investigating intrinsic patterns of functional brain connectivity have proven to be particularly compelling. Alterations in several intrinsic functional networks have been implicated in trauma-related dissociation, including the default mode network, central executive network, and salience network (Lebois, Kumar, et al., 2022; Lotfinia et al., 2020). The identification of altered connectivity within these key networks represents an important step forward in understanding the neurobiology of dissociation (Lanius et al., 2005, 2010; Lebois, Kumar, et al., 2022). However, questions remain regarding whether patterns of functional connectivity might be used to *predict* dissociative symptoms, rather than simply observing associations.

Recent work has harnessed the power of machine learning in service of identifying a neural signature that can predict severe pathological dissociation (Lebois et al., 2021; Nicholson et al., 2019; Reinders et al., 2019). Of particular relevance to the present investigation, our group developed a model to predict dissociation from intrinsic patterns of functional connectivity in a transdiagnostic group of patients with trauma-related dissociation (Lebois et al., 2021). An important next step in elaborating and refining the neural signature of pathological dissociation is to understand factors related to the performance of the predictive model. For example, it remains unclear whether clinically-relevant factors may be associated with model performance, such as diagnosis or symptom severity. Exploring the nuances of the predictive model will allow for continued refinement and ensure appropriate application of the model. Therefore, the present study aimed to investigate clinical patterns in our previously-published model predicting self-reported dissociation from intrinsic patterns of brain functional connectivity. We conducted k-means clustering to determine the presence of discrete groups in the sample and then explored group differences in a range of clinically relevant variables, such as distribution of diagnosis groups, PTSD severity, and childhood trauma severity.

## Methods

### Participants

The present work is a secondary data analysis of a sample that has been previously described (Lebois et al., 2021). Briefly, participants were 65 women seeking treatment across several levels of care (inpatient, partial hospital, and outpatient) at a psychiatric hospital in the northeastern United States. Participants had an average age of 34.4 years (*SD* = 12.2) and all participants reported a history of childhood maltreatment, current symptoms of PTSD, and varying levels of dissociation, including some who met criteria for DID (Lebois et al., 2021). Table 1 presents demographic and sample information across diagnostic groups (i.e., PTSD, PTSD dissociative subtype, and DID). All procedures of the initial study were approved by the Mass General Brigham Human Research Affairs Institutional Review Board and all study procedures were completed in accordance with the Declaration of Helsinki. All participants provided written informed consent.

**Table 1.**
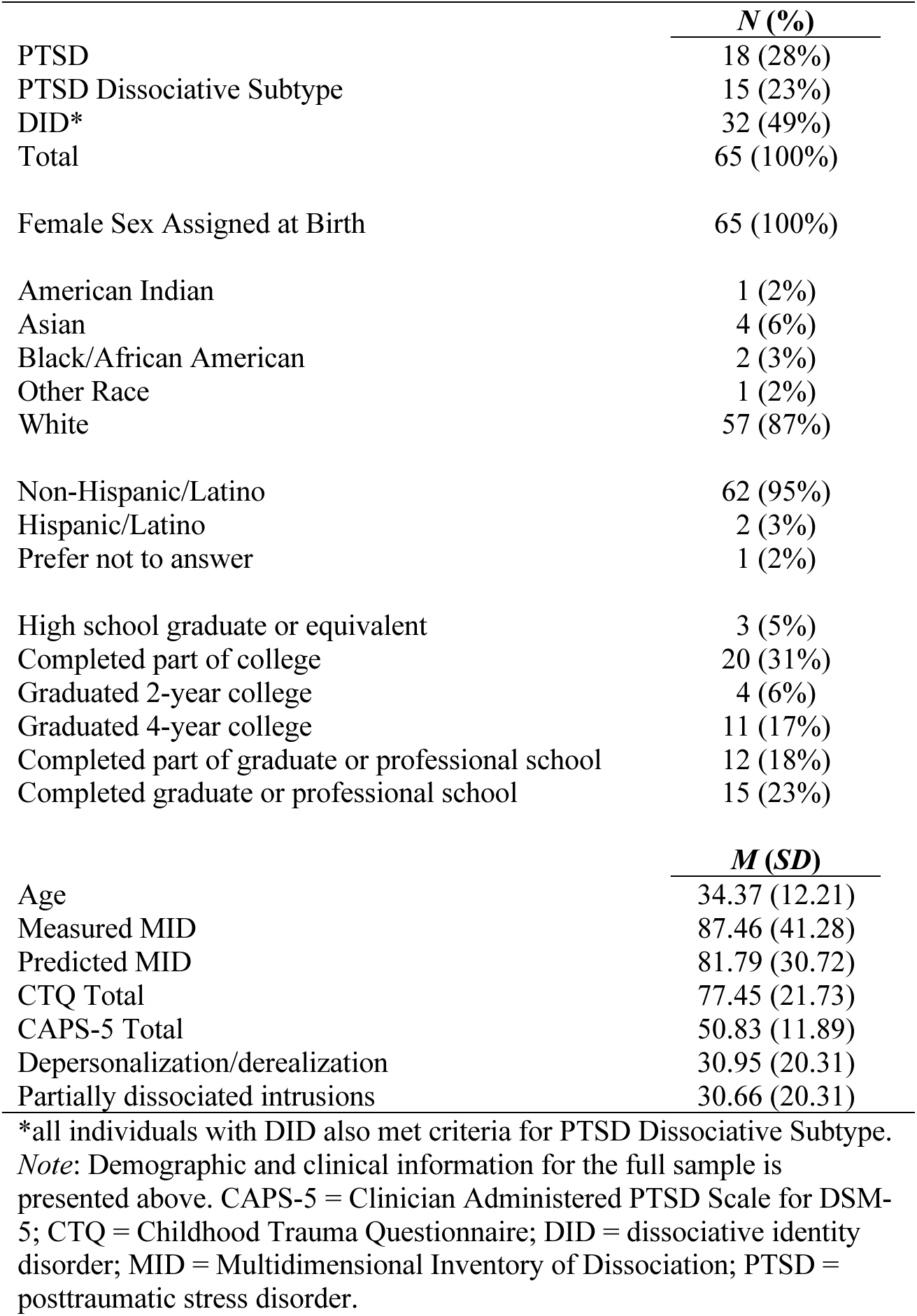
Sample demographic information.

### Measures

#### Structured Clinical Interview for DSM-IV Dissociative Disorders Revised (SCID-D-R) (Steinberg, 1994)

The SCID-D-R is a structured, clinician-administered interview and was used to diagnose dissociative disorders in the present sample. The SCID-D assesses symptoms related to a range of dissociative experiences and is currently the gold standard for assessing and diagnosing dissociative disorders, including DID.

#### Clinician Administered PTSD Scale for DSM-5 (CAPS-5) (Weathers et al., 2013)

The CAPS-5 was completed to diagnose PTSD in the present sample. Additionally, the total CAPS-5 score was utilized as a dimensional measure of PTSD symptom severity across all symptom clusters. Possible scores range from 0 to 100 and the measure displayed good internal consistency in the present sample (Chronbach’s alpha = 0.83).

#### Multidimensional Inventory of Dissociation (MID) (Dell, 2006)

The MID is a comprehensive self-report measure assessing a wide range of dissociative experiences. The MID produces scores for several subscales, including severe pathological dissociation, depersonalization/derealization, and partially dissociated intrusions. The severe pathological dissociation subscale represents the total number of items which surpassed the clinical level of significance (possible range 0 to 168). The MID also includes subscales for average symptoms of depersonalization (a sense of disconnection from one’s own internal experience) and derealization (the experience of disconnection from the external environment).

In the present study, these two subscales were averaged to produce average an depersonalization/derealization score. Finally, partially-dissociated intrusions are a cluster of experiences in which the individual experiences the partial intruding of a dissociated self-state into their conscious awareness (Coy et al., 2022; Dell, 2009). The MID evaluates 11 types of partially-dissociated intrusion experiences, such as experiencing an internal struggle, temporary losses of knowledge, and experiences of self-alteration. In contrast to the depersonalization and derealization subscales, which are considered “General Posttraumatic Dissociative Symptoms”, partially-dissociated intrusions reflect dissociative processes that are indicative of a dissociative disorder (Coy et al., 2022; Dell, 2009). In the present study, we calculated an average partially-dissociated intrusion score by averaging responses to all items. Overall, the MID displayed excellent internal consistency in the present sample (Chronbach’s alpha = 0.99).

#### Childhood Trauma Questionnaire (CTQ) (Bernstein et al., 1994)

Participants completed the CTQ, which provides a total score representing the frequency of a range of maltreatment experiences occurring prior to the age of 18. Possible scores on the CTQ range from 25 to 125 and displayed good internal consistency in the present sample (Chronbach’s alpha = 0.94).

### Procedures

Participants in the original study completed diagnostic interviews and self-report questionnaires, as well as a MRI scanning session. These procedures have been described in detail elsewhere (Lebois et al., 2021). Briefly, all MRI sessions were completed on a 3T Siemens Trio (Siemens Healthcare) using a 12-channel head coil. Participants completed several task-based scans and a rest scan, all using standard T_2_*-weighted echo-planar imaging sequences (TR = 3000ms, TE = 30ms, flip angle = 85°, 3×3×3mm voxels) for blood-oxygen-level dependent (BOLD) fMRI. A multiecho magnetization-prepared rapid acquisition gradient echo (MEMPRAGE) anatomical scan was also completed (TR = 2530ms, TE 1 = 1.64ms, TE 2 = 3.5ms, TE 3 = 5.36ms, TE 4 = 7.22ms flip angle = 7°, 1×1×1mm voxels).

### Neuroimaging Data Preprocessing and Analysis from Original Publication

The neuroimaging analysis that produced the prediction model we examined in this paper is presented in a prior publication (Lebois et al., 2021). However, for context, we briefly describe the processing procedures next. All fMRI data (i.e., task-based and resting-state) were processed in the same way, consistent with previously published work (Buckner et al., 2011). Preprocessing included slice timing correction, motion correction, bandpass filtering, motion regression, whole-brain signal regression, and ventricular and white matter signal regression. Individually-based functional regions of interest were identified using an iterative process, described elsewhere (Li et al., 2019; Wang et al., 2015), based on the 92 regions of interest from the Genomic Superstruct Project (Holmes et al., 2015). This process resulted in 92 individually-sized functional regions of interest per participant (Lebois et al., 2021). Next, whole-brain functional connectivity matrices were estimated for each participant. Then, a support vector machine for regression (SVR) algorithm was used to identify symptom-connectivity associations in which a participant’s functional connectivity was correlated with their MID severe pathological dissociation score (Lebois et al., 2021). Covariates included in the model were measures of childhood maltreatment (CTQ total score), PTSD symptom severity (CAPS-5 total score), participant age, and measures of head motion inside the scanner. The SVR model then utilized the identified functional connections to predict MID severe dissociation score (Lebois et al., 2021). The present study used the measured (i.e., self-reported) and predicted (i.e., estimated based on brain functional connectivity) MID scores generated by the prediction SVR model.

### Statistical Analyses

All analyses were completed in R (version 4.3.1) (R Core Team, 2023). First, we conducted a cluster analysis (*kmeans*) to explore patterns in the previously-identified prediction model. We used the measured and predicted MID scores from the original predictive model (Lebois et al., 2021) as the input to the clustering algorithm. We tested cluster sizes ranging from k=2 to k=5 and determined the optimal number of clusters for the present data by visualizing a scree plot (*kmeans* method = “wss”) and a silhouette plot (*kmeans* method = “silhouette”). We examined group differences among the identified clusters using chi-square tests and ANOVAs. Specifically, we explored whether the groups differed in a range of clinically relevant variables, including diagnosis distribution, predicted and measured severe pathological dissociation MID scores, overall PTSD severity (CAPS-5), experiences of childhood trauma (CTQ), MID depersonalization/derealization, and MID partially dissociated intrusions. Overall ANOVAs were Bonferroni corrected (new ANOVA critical *p*-value = 0.05/6 = 0.008). In cases where the overall ANOVA was significant, we followed up with post-hoc t-tests and used Tukey HSD (*TukeyHSD*) for multiple comparison correction in R.

## Results

### Clustering Results

Descriptive information for the overall sample can be found in Table 1. Both the scree and silhouette plots indicated the presence of four distinct clusters in the present data (Figure 1). A visual depiction of the four-cluster solution can be found in Figure 2 and descriptive information for each of the four clusters can be found in Table 2. Additionally, we computed the mean squared error (MSE) for each cluster (measured MID – predicted MID), which serves as an index of accuracy in the original predictive model. Lower MSE values reflect less discrepancy between measured MID and predicted MID, thus reflecting greater accuracy. Cluster 1 had the lowest MSE (749.69), followed by cluster 4 (900.56), cluster 3 (1704.17), and cluster 2 had the highest MSE (2827.44).

**Figure 1.**
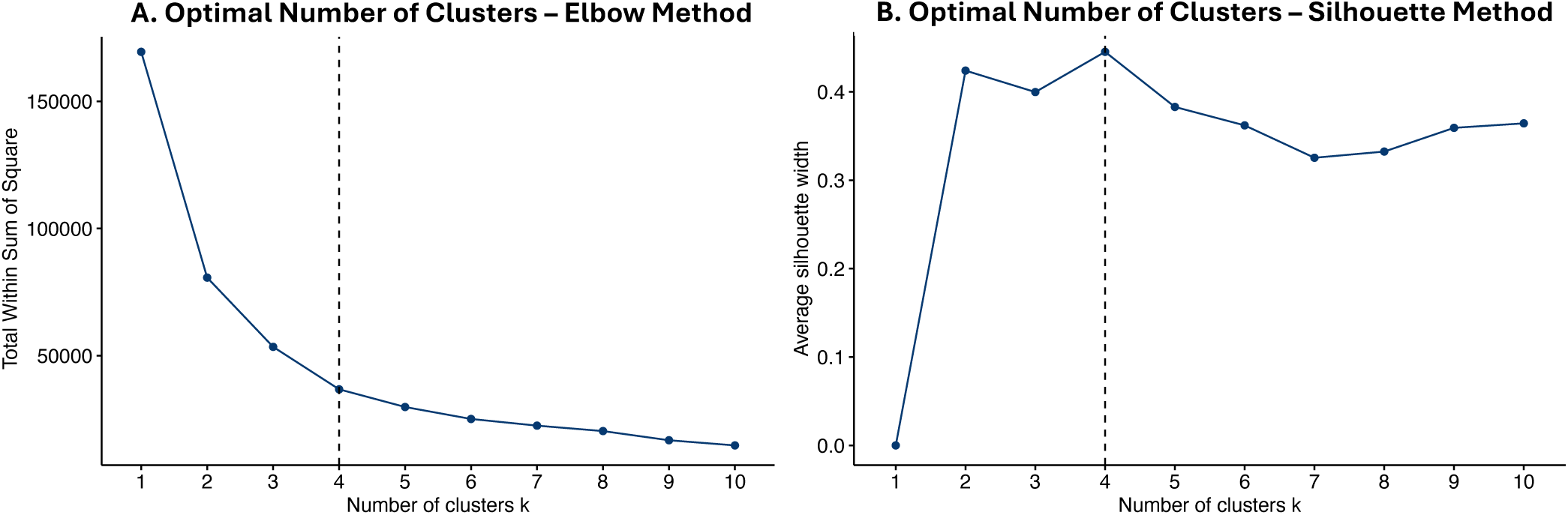
Plots showing the optimal number of clusters in the present data. Panel A shows a scree plot using the elbow method, while Panel B shows a plot using the silhouette method. The vertical dashed lines in both Panels A and B show the optimal number of clusters, which was found to be k=4 for both methods.

**Figure 2.**
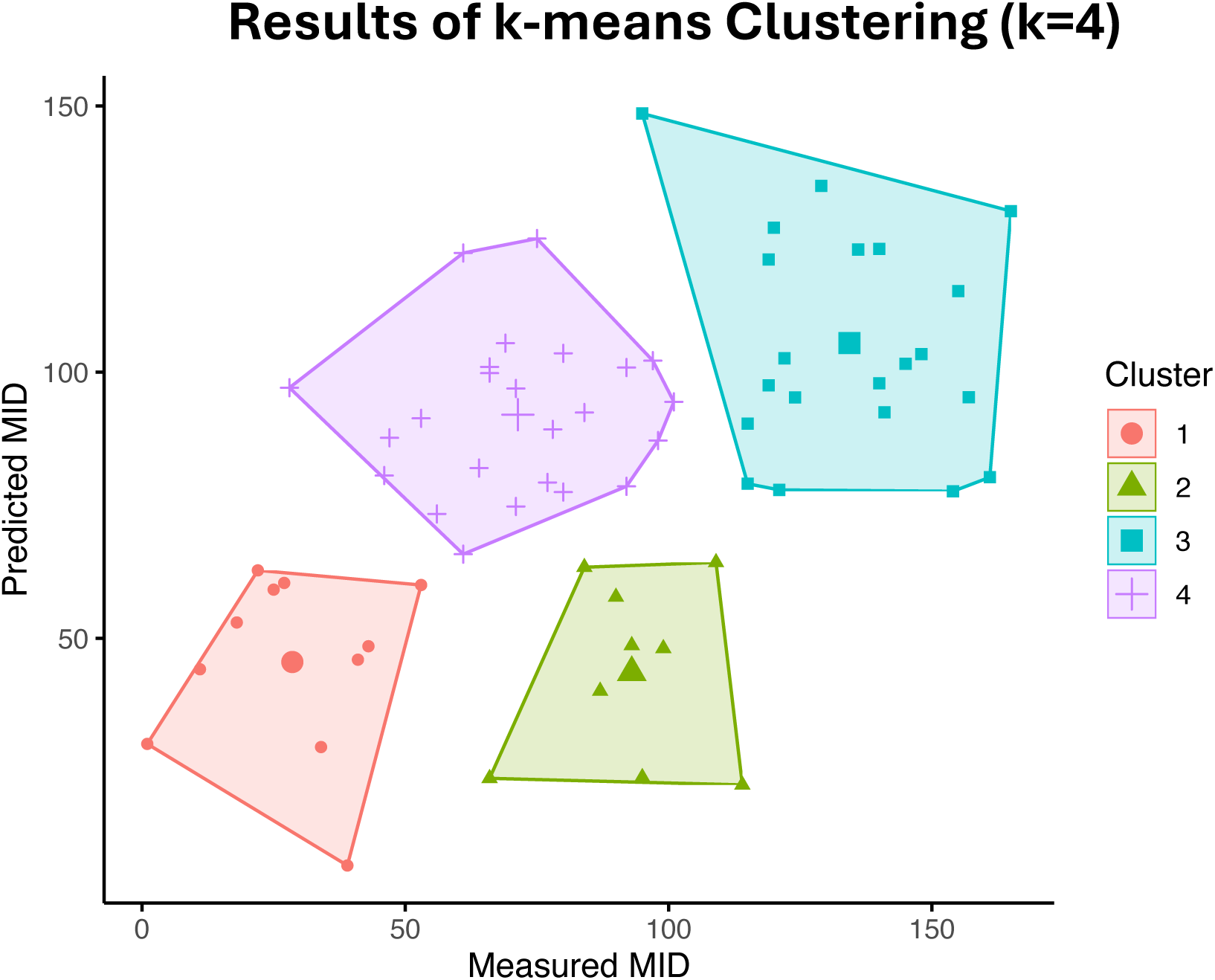
Graphical depiction of the four clusters found in the present data. Clusters are identified both by color and icon shape, as shown in the legend. Smaller icons represent individual participants, while the larger icons represent the centroid of each cluster. Measured MID (x-axis) and predicted MID (y-axis) were collected as part of the original study from which these data were drawn (Lebois et al., 2021). Specifically, predicted MID refers to the MID score estimated on the basis of brain functional connectivity. Please refer to the online version of the manuscript for a color figure.

**Table 2.** Cluster-specific clinical information.

|  | **Cluster 1**<br>(*n* = 11) | **Cluster 2**<br>(*n* = 9) | **Cluster 3**<br>(*n* = 21) | **Cluster 4**<br>(*n* = 24) |
| --- | --- | --- | --- | --- |
|  | ***n* (%)** | ***n* (%)** | ***n* (%)** | ***n* (%)** |
| PTSD | 9 (82%) | 1 (11%) | 0 (0%) | 8 (33%) |
| PTSD Dissociative Subtype | 2 (18%) | 2 (22%) | 3 (14%) | 8 (33%) |
| DID | 0 (0%) | 6 (67%) | 18 (86%) | 8 (33%) |
|  | ***M* (*SD*)** | ***M* (*SD*)** | ***M* (*SD*)** | ***M* (*SD*)** |
| Measured MID | 28.55 (15.29) | 93.00 (14.11) | 143.33 (18.48) | 71.38 (18.08) |
| Predicted MID | 44.55 (17.12) | 43.59 (16.98) | 105.47 (20.50) | 92.02 (14.56) |
| CTQ Total | 55.55 (15.23) | 66.11 (24.43) | 87.29 (10.94) | 83.12 (22.36) |
| CAPS-5 Total | 44.18 (9.14) | 42.56 (10.55) | 56.62 (11.47) | 51.92 (11.14) |
| Depersonalization/derealization MID | 8.90 (8.07) | 30.46 (15.80) | 49.36 (17.58) | 25.12 (13.65) |
| Partially-dissociated intrusions MID | 8.77 (5.01) | 31.28 (12.50) | 53.85 (13.01) | 20.16 (10.01) |

*Note:* Clinical information for each of the four clusters is presented above. Comparisons among the four clusters are depicted in Figure 4. CAPS-5 = Clinician Administered PTSD Scale for DSM-5; CTQ = Childhood Trauma Questionnaire; DID = dissociative identity disorder; MID = Multidimensional Inventory of Dissociation; PTSD = posttraumatic stress disorder.

### Diagnostic Breakdown between Identified Clusters

Chi-square tests indicated that the distribution of diagnoses differed across clusters [*χ*^2^(6) = 33.42; *p* < 0.001; Figure 3]. Cluster 1 (n=11) was comprised primarily of PTSD patients (82%) with a smaller proportion of dissociative subtype patients (18%) and no DID patients. Cluster 2 (n=9) was made up of a small majority of DID patients (67%), with a smaller proportion of PTSD dissociative subtype patients (22%) and a minority of PTSD patients (11%). Cluster 3 (n=21) was overwhelmingly comprised of DID patients (86%), with a small proportion of dissociative subtype patients (14%) and no PTSD patients. Finally, cluster 4 (n=24) was evenly split, with each diagnosis making up one third of the total group.

**Figure 3.**
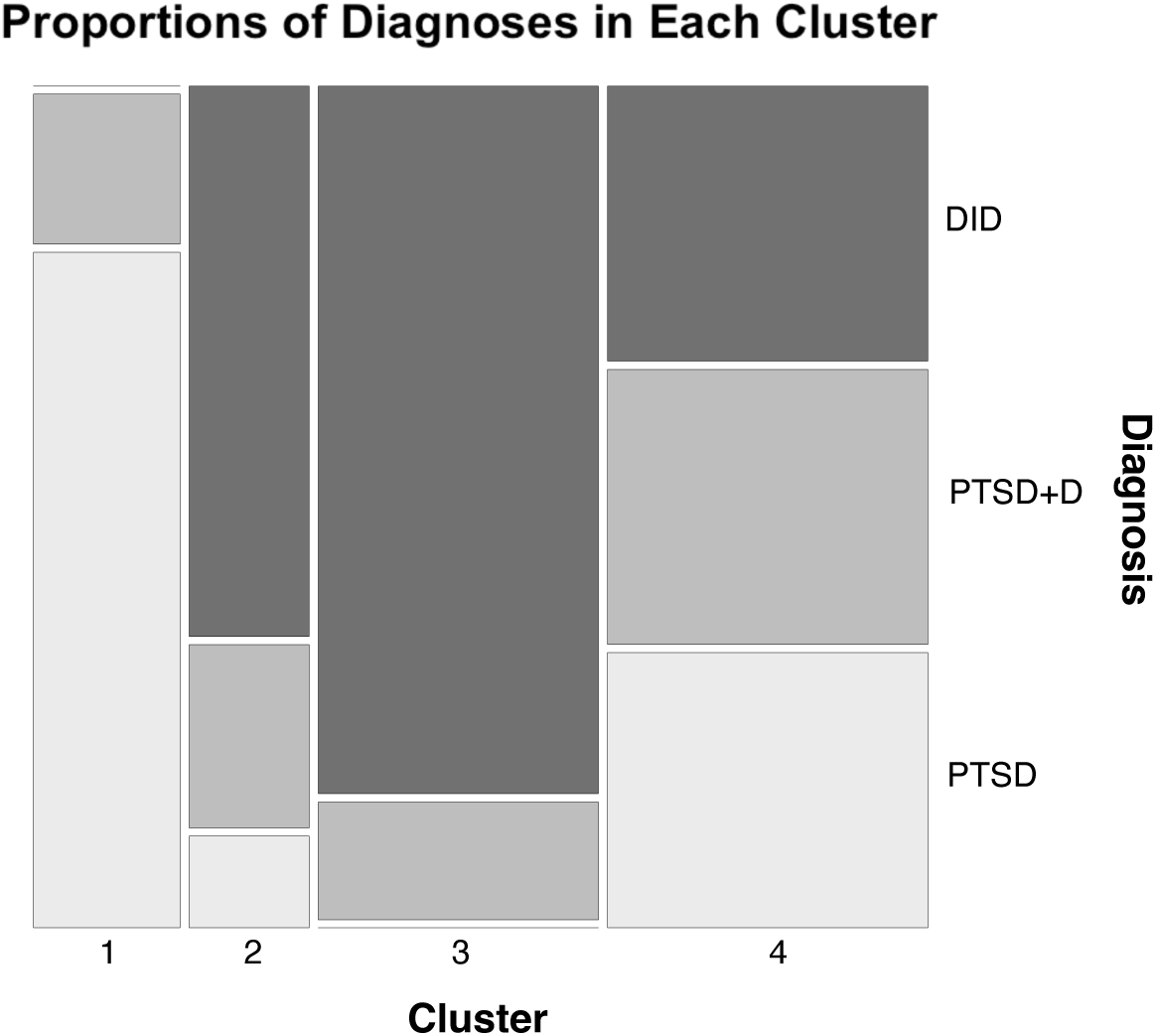
Mosaic plot showing the distribution of diagnoses across the four clusters. The darkest shaded boxes represent individuals with dissociative identity disorder (DID). Medium shaded boxes represent individuals with the dissociative subtype of posttraumatic stress disorder (PTSD+D). The lightest shaded boxes represent individuals with posttraumatic stress disorder (PTSD). Solid light gray lines indicate that there were no participants with that diagnosis in a particular cluster. Specifically, there were no participants with DID in Cluster 1 and no participants with the dissociative subtype of PTSD in Cluster 3.

### Differences in Dimensional Symptoms across Identified Clusters

As expected, ANOVAs revealed significant differences in measured MID [*F*(3) = 101.00, *p* < 0.001] and predicted MID scores [*F*(3) = 45.96, *p* < 0.001], which further support the validity of the 4-group solution. Additionally, ANOVAs revealed significant group differences in clinically-relevant variables including CTQ score [*F*(3) = 8.95, *p* < 0.001], PTSD severity [*F*(3) = 5.17, *p* = 0.003], depersonalization/derealization [*F*(3) = 20.60, *p* < 0.001], and partially dissociated intrusions [*F*(3) = 54.33, *p* < 0.001]. Results of Tukey-corrected post-hoc tests are shown in Figure 4. On the CTQ, cluster 1 had a lower score than clusters 3 and 4, while cluster 2 had a lower score than cluster 3. Both clusters 1 and 2 again had lower PTSD severity than cluster 3 but did not differ from each other. In terms of dissociation-specific symptoms, a similar pattern emerged for both depersonalization/derealization and partially-dissociation intrusions. In general, cluster 1 had the lowest scores on both measures while clusters 2 and 4 fell in the middle and did not differ from each other. Cluster 3 had the highest scores on both measures and differed from both clusters 2 and 4. Cluster-specific descriptive information can be found in Table 2 and full results of post-hoc tests can be found in Figure 4.

**Figure 4.**
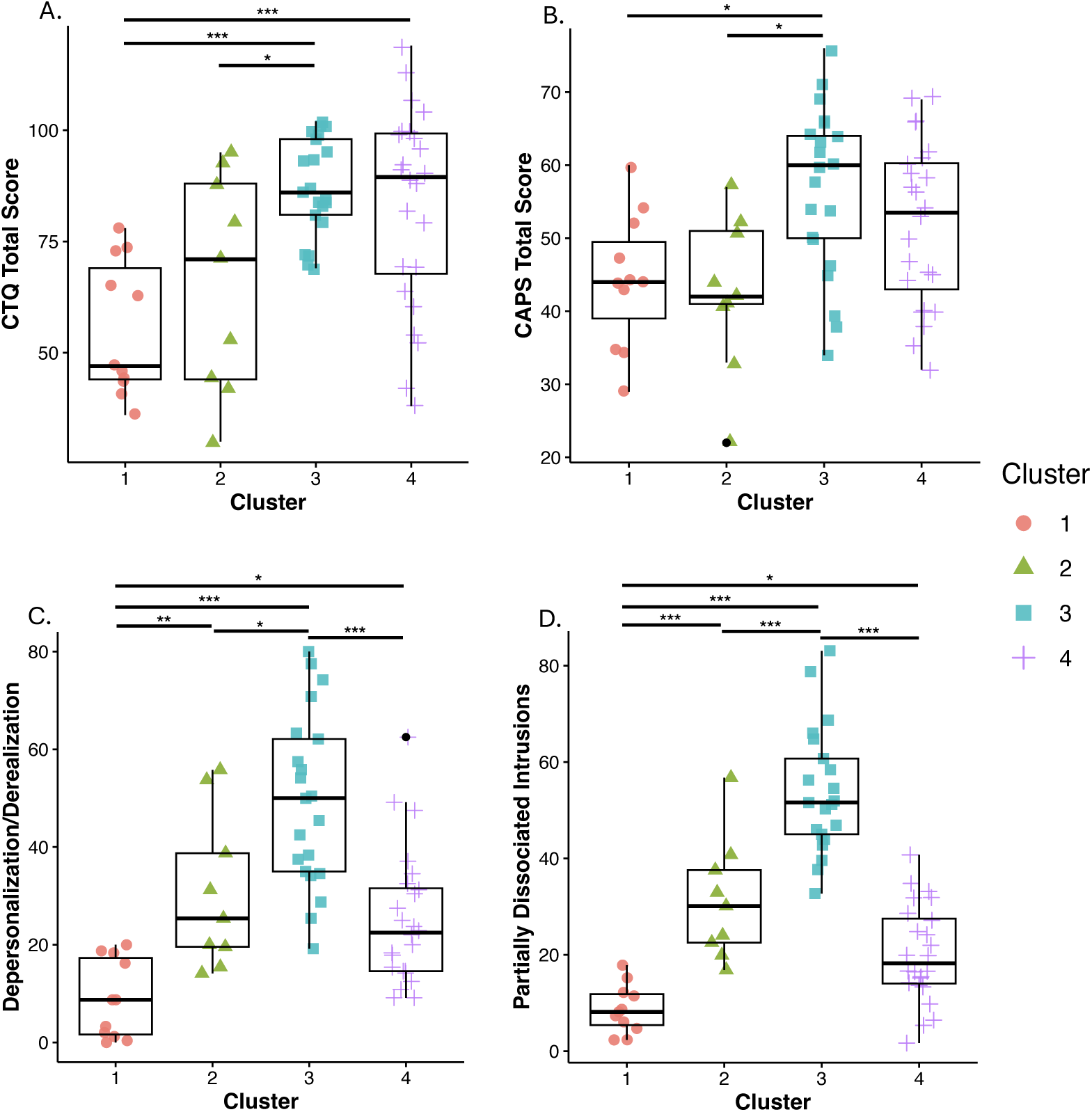
Boxplots showing the results of post-hoc tests examining differences among the four clusters on clinically-relevant variables. (A) childhood maltreatment (CTQ total score); (B) PTSD severity (CAPS-5 total score); (C) depersonalization/derealization; (D) partially-dissociated intrusions. Horizontal bars indicate significant differences. \**p* < 0.05; \*\**p* < 0.01; *** *p* < 0.001.

### Exploratory Post-hoc analysis of Participants with DID in Cluster 2 and 3

Clusters 2 and 3 were both comprised primarily of individuals with DID (Figure 3), though they differed in all clinically-relevant variables we investigated. Therefore, we conducted exploratory post-hoc analyses to examine whether participants with DID in cluster 2 differed from individuals with DID in cluster 3 on the same clinical variables described above (i.e., measured and predicted MID, CTQ, PTSD severity, depersonalization/derealization, and partially-dissociated intrusions). We again used the Bonferroni-corrected critical *p*-value of 0.008. Compared to participants with DID in cluster 2, those with DID in cluster 3 had higher scores on the measured MID, predicted MID, PTSD symptom severity, and partially dissociated intrusions. The two groups did not differ in CTQ score or depersonalization/derealization. Full results can be found in Table 3 and Figure 5. Finally, when considering only participants with DID, the MSE of cluster 2 was 3505.36 and the MSE of cluster 3 was 1756.99.

**Figure 5.**
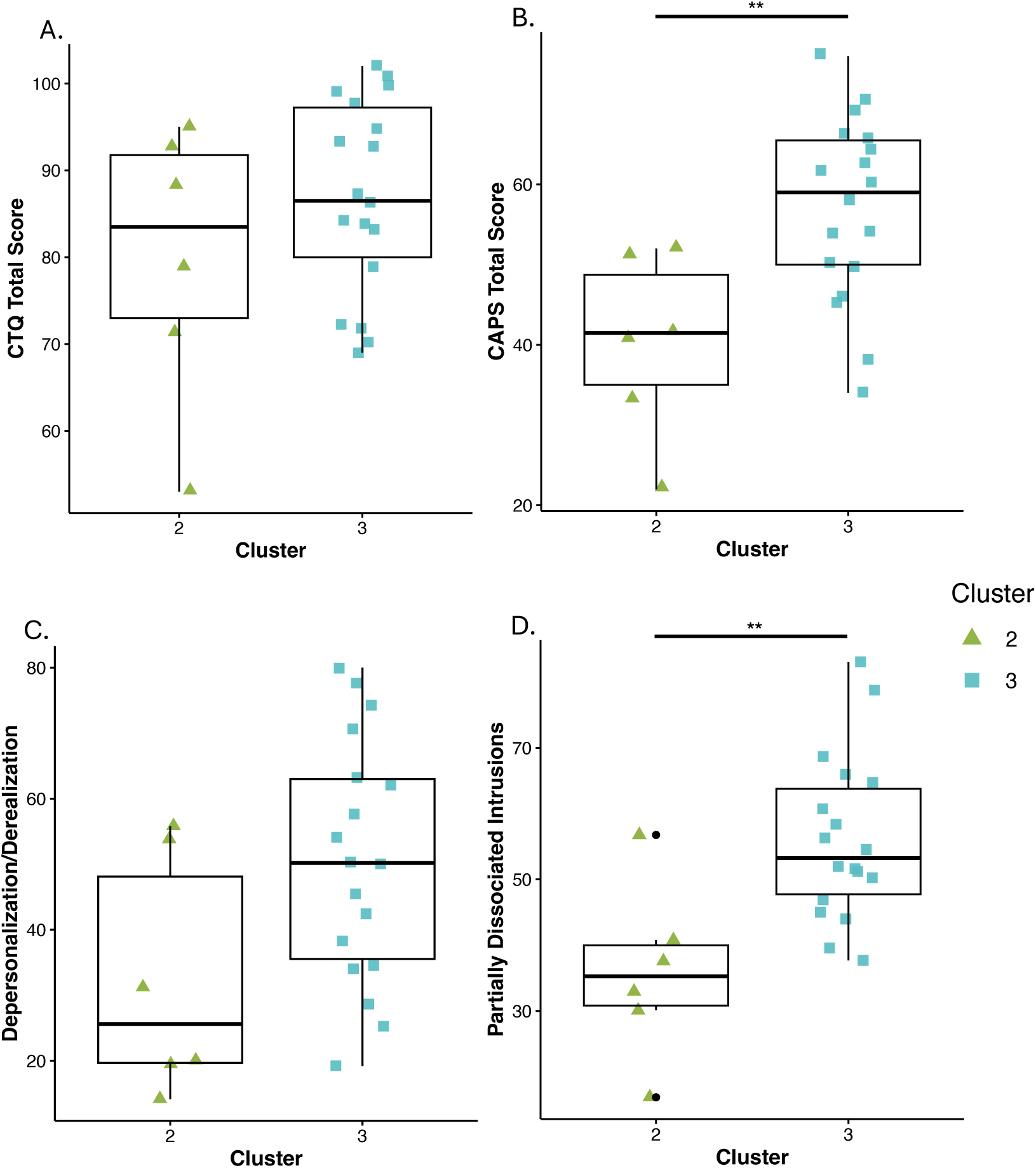
Boxplots showing the results of exploratory analyses examining differences between participants with DID in clusters 2 and 3. (A) childhood maltreatment (CTQ total score); (B) PTSD severity (CAPS-5 total score); (C) depersonalization/derealization; (D) partially-dissociated intrusions. Horizontal bars indicate significant differences. \*\**p* < 0.01

**Table 3.** Clinical information for exploratory follow up analyses.

|  | Cluster 2 | Cluster 3 |  |
| --- | --- | --- | --- |
| Number of DID participants | 6 | 18 |  |
|  | *M (SD)* | *M (SD)* | ANOVA |
| Measured MID | 100.00 (9.51) | 137.94 (16.63) | ***F(1) = 27.65, p < 0.001*** |
| Predicted MID | 44.19 (17.36) | 104.08 (19.31) | ***F(1) = 45.24, p < 0.001*** |
| CTQ Total | 79.83 (15.93) | 87.06 (11.25) | *F(1) = 1.51, p = 0.232* |
| CAPS-5 Total | 40.17 (11.34) | 57.00 (11.51) | ***F(1) = 9.684, p = 0.005*** |
| Depersonalization/derealization MID | 32.42 (18.20) | 50.46 (18.43) | *F(1) = 4.336, p = 0.05* |
| Partially dissociated intrusions MID | 35.84 (13.16) | 56.08 (12.52) | ***F(1) = 11.49, p = 0.003*** |

*Note:* Clinical information only for participants with DID in Clusters 2 and 3 is presented above. ANOVAs were conducted to compare the two groups on each of the clinical variables. Bolded values indicate significant tests. CAPS-5 = Clinician Administered PTSD Scale for DSM-5; CTQ = Childhood Trauma Questionnaire; DID = dissociative identity disorder; MID = Multidimensional Inventory of Dissociation; PTSD = posttraumatic stress disorder.

## Discussion

Previously published work developed a machine learning model using patterns of brain functional connectivity to predict symptoms of dissociation in a transdiagnostic group of women. An important next step in model development is the exploration of factors associated with performance of the predictive model. Therefore, we conducted a k-means clustering analysis and examined differences among the identified clusters on a range of clinically-relevant variables. Exploring patterns in the original predictive model will provide essential information to support future model refinement and elaboration.

Our analyses identified four discrete clusters of participants with distinct diagnostic and symptom profiles. Results of the clustering analysis were striking given our relatively homogenous sample of trauma-exposed, treatment-seeking, predominantly white adult women. Importantly, the cluster-level symptom profiles were consistent with patterns that would be expected based on diagnostic profiles of each cluster. For example, prior work has consistently identified a distinct symptom profile for the dissociative subtype of PTSD (Hansen et al., 2017; Misitano et al., 2024; Wolf, Lunney, et al., 2012; Wolf, Miller, et al., 2012). Fewer investigations have examined complex dissociative disorders, though our results are in line with the existing literature. Specifically, prior work indicates that participants with DID generally report higher levels of childhood abuse and neglect, as well as more severe symptoms of PTSD, than individuals with PTSD alone (Brand et al., 2009, 2016; Lebois, Kumar, et al., 2022; Purcell et al., 2024). Similarly, we found that the cluster with the largest proportion of participants with DID (cluster 3) reported greater childhood abuse and neglect and current PTSD symptom severity than cluster 1, which was comprised predominantly of participants with PTSD alone. Unexpectedly, participants in clusters 2 and 3 demonstrated different symptom profiles although both groups included primarily participants with DID. Therefore, we further investigated these clusters to better understand the observed divergent symptom profiles.

We identified key differences between participants with DID in clusters 2 and 3. Participants differed both in PTSD symptom severity and partially-dissociated intrusions, a type of dissociative experience that is specific to DID (Coy et al., 2022; Dell, 2006; Lebois, Kumar, et al., 2022). Partially-dissociated intrusions represent an alteration in a cohesive sense of self in which an individual experiences their thoughts, emotions, or body sensations as “intruding” into their conscious awareness, along with a lack of felt ownership of the experience (Dell, 2006; Lebois, Kumar, et al., 2022). Reducing the frequency and intensity of these partially-dissociated intrusions is a key goal of psychotherapy in DID and is accomplished by slowly, gradually supporting the individual in building the capacity to experience their trauma-related thoughts, feelings, and memories as their own (International Society For The Study of Trauma and Dissociation, 2011; Purcell et al., 2024). The term integration is often used to describe this extended process and the resulting decrease in the experience of partially-dissociated intrusions (International Society For The Study of Trauma and Dissociation, 2011). These findings suggest that partially-dissociated intrusions, and variation in integration between individuals, may represent an important and overlooked source of variability in studies of individuals with complex dissociative disorders, such as DID. This particular facet of dissociation frequently goes unmeasured in research investigations, given that it is not comprehensively captured by the most commonly used symptom screening tool, the Dissociative Experiences Scale (Brand et al., 2006; Brand & Loewenstein, 2010; Carlson & Putnam, 1993). Further, our results suggest that variability in the severity of partially-dissociated intrusions among participants with DID may influence performance of the original predictive model. We found that the predictive model was most accurate for participants in cluster 1, who reported relatively low levels of dissociation, and least accurate for participants in cluster 2, who reported low levels of partially-dissociated intrusions. In fact, the MSE of cluster 2 was even larger when considering only participants with DID (3505.36 vs 2827.44), highlighting that diminished accuracy of the predictive model was likely driven by the participants with DID in this cluster. These findings underscore the importance of utilizing comprehensive assessments of dissociation whenever possible, as well as the necessity of thoroughly understanding factors impacting predictive model performance.

Results of the present investigation revealed several important avenues for future work to refine and extend the original predictive model. The original model predicted dissociative symptoms on the basis of patterns of brain functional connectivity, which we have not examined here. Given that we identified distinct participant groups in the behavioral data, an important next step will be to understand possible group differences in the patterns of functional connectivity. Additionally, the sample in which the original predictive model was developed was relatively small and homogenous, thus it remains unclear to what extent our findings may generalize. Replication studies in larger and more diverse samples will be necessary to further validate the predictive model. Continued model refinement is essential to enhance predictive accuracy and to identify appropriate circumstances under which the model should be implemented. For example, the original predictive model was developed using a sample of participants with an average age of 34. However, patterns of functional connectivity shift across the lifespan (Casey et al., 2016; Casey & Jones, 2010). Thus, at present, the model may not be as accurate in children, younger adults, or older adults, as the impact of age-related shifts in functional connectivity patterns remains unclear.

We investigated patterns in a previously-identified machine learning model which predicted dissociative symptoms on the basis of brain functional connectivity in a transdiagnostic sample of adult women. Our analyses identified four discrete participant clusters, with distinct diagnostic and symptom profiles. Further, we found that DID-specific types of dissociation, such as partially-dissociated intrusions, represent an important and often overlooked source of variability in research. Findings of the present investigation highlight the need to thoroughly investigate factors impacting performance of machine learning models. The development of the original predictive model represented an exciting step toward precision medicine and the identification of objective indices of dissociative experiences. Future work centered on model refinement and elaboration will continue the progress toward achieving these goals. Our hope is that the elaboration of neurobiological signatures of dissociation will contribute to better clinical understanding of the condition, stimulate development of novel treatment targets, and, ultimately, reduce the pain and suffering of those living with dissociation.

## Data Availability

Participant data used in the present study are available upon reasonable request to the authors and approval by appropriate regulatory bodies.

## Notes

### Competing Interest Statement

LAM Lebois has an unpaid membership on the Scientific Committee for the International Society for the Study of Trauma and Dissociation (ISSTD). They also report spousal IP payments from Vanderbilt University for technology licensed to Acadia Pharmaceuticals and spousal equity/ownership interest in Violet Therapeutics unrelated to the present work. ML Kaufman, CA Palermo, and M Robinson report unpaid membership on the Scientific Committee for the ISSTD. The authors have no other relevant affiliations or financial involvement with any organization or entity with a financial interest in or financial conflict with the subject matter or materials discussed in the manuscript apart from those disclosed.

### Funding Statement

JB Purcell, CA Palermo, ZA Bair, RL Modell, MA Robinson, ML Kaufman, and LAM Lebois are funded by the Julia Kasparian Fund for Neuroscience Research and by the National Institute of Mental Health, the United States via grants K01 MH118467 and R01 MH119227. MC Marr is funded by the National Institute of Mental Health via grant R25 MH13587. This work was conducted with support from UM1TR004408 award through Harvard Catalyst | The Harvard Clinical and Translational Science Center (National Center for Advancing Translational Sciences, National Institutes of Health) and financial contributions from Harvard University and its affiliated academic healthcare centers. The content is solely the responsibility of the authors and does not necessarily represent the official views of Harvard Catalyst, Harvard University and its affiliated academic healthcare centers, or the National Institutes of Health. Neither ISSTD nor any funding sources were involved in the analysis or preparation of the paper.

### Author Declarations

The Institutional Review Board of Mass General Brigham Human Research Affairs gave ethical approval for this work.

